# Functional Brain Age Acceleration from Dynamic and Static Connectivity Predicts Working Memory and Attention Deficits in Schizophrenia

**DOI:** 10.1101/2025.07.21.25331956

**Authors:** Sabrina J. Edwards-Swart, Bradley Baker, Daniel H. Mathalon, Judith M. Ford, Adrian Preda, Theo G.M. van Erp, Godfrey D. Pearlson, Jessica A. Turner, Vince D. Calhoun, Mohammad S. E. Sendi

## Abstract

**Background:** Schizophrenia is characterized by deficits in attention and working memory. In recent years, the brain age gap (BAG), defined as the difference between neuroimaging-predicted and chronological age, has emerged as a biomarker of brain dysfunction. Prior studies primarily use structural MRI or static functional network connectivity (sFNC), while the potential of dynamic functional network connectivity (dFNC) to quantify BAG in relationship with cognition remains underexplored.

**Methods:** Leveraging one of the largest resting-state fMRI datasets to date (N=22,569; UK Biobank, HCP, HCP-Aging), we developed robust brain age prediction models incorporating both wide-brain and sub-network variables derived from sFNC and dFNC. These models were validated in an independent clinical sample (FBIRN; N=153) including individuals with schizophrenia and healthy controls. Associations between BAGs and cognitive measures (attention vigilance, working memory) were evaluated using general linear models, controlling for key demographic and clinical covariates.

**Results:** Both sFNC and dFNC models demonstrated robust prediction accuracy in healthy individuals (sFNC: *r*=0.8755; dFNC: *r*=0.8675). Wide-brain BAGs (wBAGs) showed strong negative associations with attention vigilance (sFNC: *r*=–0.2923, *FDR p*=0.0013; dFNC: *r*=– 0.2715, *FDR p*=0.0016) and working memory (sFNC: *r*=–0.2237, *FDR p*=0.0088; dFNC: *r*=– 0.2508, *FDR p*=0.0064). Sub-network BAGs (subBAGs) within subcortical, sensorimotor, cognitive control, and default mode networks robustly predicted cognitive deficits, with dFNC-derived subBAGs showing the strongest effects (*FDR p*<0.01).

**Conclusions:** Our findings establish dFNC-based BAGs as a sensitive and clinically relevant biomarker of cognitive impairment in schizophrenia, outperforming sFNC and highlighting the translational potential of dynamic connectivity for precision diagnosis and treatment.

## 1. INTRODUCTION

Schizophrenia, affecting 0.25% to 0.64% of the U.S. population, is a complex psychiatric disorder marked by profound disruptions in cognition, emotion, and social interactions (1,2). Key symptoms include hallucinations, delusions, and motor impairments, alongside significant reductions in emotional expression and social engagement. Critically, individuals with schizophrenia exhibit marked cognitive deficits, particularly in working memory and attention vigilance. Studies indicate they perform approximately 2.5 standard deviations below healthy controls in these cognitive domains (3). These deficits are characterized by difficulties in retaining and updating information in working memory (4), with increasing cognitive demands leading to more pronounced declines in attention vigilance compared to healthy individuals (5).

In recent decades, neuroimaging techniques have greatly enhanced our understanding of cognitive function and attention deficits in schizophrenia (4,6). Structural magnetic resonance imaging (sMRI) provides detailed anatomical insights (7), revealed associations between executive function impairments and reductions in prefrontal cortex volume and thickness, and linked episodic memory deficits with hippocampal atrophy (8). Conversely, functional magnetic resonance imaging (fMRI) measures brain activity through oxygen metabolism and blood flow, illuminated functional dynamics across various brain regions (9). Functional MRI has been pivotal in identifying brain regions involved in working memory, offering valuable insights into cognitive impairments in patient populations compared to healthy controls. Broadly, Neuroimaging plays a key role in examining executive function and episodic memory impairments and provides a valuable tool for tracking brain function changes following cognitive remediation therapies (8).

In recent years, brain age prediction from neuroimaging has emerged as a promising approach for identifying novel biomarkers of neurological and neuropsychiatric conditions (14,24,26,47,49,51). By modeling the relationship between neuroimaging variables and age in healthy individuals, this method enables the detection of meaningful brain deviations in patient populations. The difference between an individual’s predicted brain age and their chronological age, termed the brain age gap (BAG), provides critical insights into the pathology of cognitive function (10–13). For example, emerging evidence demonstrates that individuals with accelerated brain age consistently show marked deficits across critical cognitive domains— including IQ, verbal comprehension, perceptual reasoning, processing speed, working memory, and memory recall—as captured by gold-standard assessments such as the Rey Auditory Verbal Learning Test (12). In schizophrenia, studies using sMRI have revealed that patients often exhibit a brain age 6 to 8 years older than their chronological age (15). Further, the BAG was noted to increase across at-risk, recent onset (16), and recurrent phases of schizophrenia, initially rising approximately a year and a half annually for the first five years before stabilizing (17). Additionally, early-stage schizophrenia and bipolar disorder present distinct BAG profiles.

Individuals with schizophrenia consistently exhibit elevated BAGs, whereas those with bipolar disorder maintain levels comparable to healthy controls. This distinction underscores the potential of BAG as a valuable biomarker for enhancing early differential diagnosis (18). This suggests a progressive pathogenic component unique to schizophrenia, underscoring the utility of BAG as a biomarker (19). Moreover, a significant negative association between BAG and cognitive functions like working memory has been observed in schizophrenia, highlighting the potential of BAG in understanding and diagnosing declined neurocognitive performance in schizophrenia (20).

While the majority of brain age prediction research has traditionally focused on structural MRI (sMRI), recent advancements include employing functional connectivity metrics from resting-state fMRI to predict brain age (21–24). A notable study using resting-state fMRI data from the Philadelphia Neurodevelopmental Cohort demonstrated the value of brain age models in youth by linking older estimated brain age to greater symptom burden, especially across DSM-5 psychiatric diagnoses. These findings underscore the potential of brain age to capture deviations in brain maturation associated with mental health conditions (25). Furthermore, an innovative study used resting-state fMRI and brain age prediction to identify neural connections associated with abnormal brain aging. By systematically excluding connections from the training model, the approach pinpointed those most critical to brain age accuracy, providing new insights into the neurobiological mechanisms of age-related psychiatric conditions (26).

Although brain age prediction using static functional connectivity (sFNC), estimated from resting-state fMRI, has proven valuable, the potential of dynamic functional connectivity (dFNC) in this area is still underexplored. Unlike sFNC, which evaluates connectivity from correlations across an entire time series, dFNC analyzes connections between brain regions or networks within specific time intervals. This allows for the capture of temporal fluctuations that reveal how brain network interactions change over time (27,28). The variability and dynamism of neural signals, captured by dFNC, are crucial in understanding cognitive deficits and clinical symptoms in psychiatric disorders. These dynamic measures provide insights into brain function that static approaches cannot, highlighting changes and interactions that are critical in disease progression (29–35). Because cognitive symptoms in schizophrenia are potentially driven by disrupted brain network dynamics, leveraging a BAG derived from dFNC offers a powerful, next-generation marker of disease-related neurobiological aging. Critically, no study has yet evaluated whether dFNC-based BAG outperforms traditional static FNC (sFNC) BAG in explaining core cognitive deficits such as working memory and attention. Establishing this added predictive power would mark a transformative advance—positioning dFNC BAG as a mechanistically grounded biomarker and unlocking new precision targets for circuit-based interventions in schizophrenia and related psychiatric conditions.

Here we address this gap using the largest sample to date: 22,569 resting-state scans from 17□522 healthy adults from UK Biobank (36,37) and Human Connectome Project (38–40) to train multiple wide-brain and sub-network brain age prediction models, and an independent cohort of 153 controls and individuals with schizophrenia from the FBIRN consortium to test them (41). Leveraging a graph-convolutional network for sFNC and a bidirectional LSTM for dFNC, we generated wide-brain and sub-network BAGs and evaluate their links to attention vigilance and working memory performance while controlling for demographic and clinical covariates. We hypothesize that (1) BAGs derived from dFNC will predict age as accurately as sFNC models, and (2) larger BAGs—particularly those based on dFNC—will associate with poorer cognitive performance, reflecting accelerated functional brain ageing in schizophrenia. By integrating dynamic connectivity with brain age modeling, this study provides the first direct evidence linking aberrant network dynamics to cognitive impairment and establishes dFNC-based BAG as a clinically relevant biomarker.

## 2. METHODS

### 2.1 Study Population

Our study leveraged multiple datasets, including the UK Biobank or UKBB (36,37) and the Human Connectome Project’s Young Adult or HCP-YA (38) and Aging or HCP-A (40) cohorts to develop brain age prediction models. For the UKBB dataset, exclusion criteria encompassed a range of mental and behavioral disorders as categorized by the International Classification of Diseases version 10 (ICD-10): delirium not induced by alcohol and other psychoactive substances (F05); mental disorders due to brain damage, dysfunction, or physical disease (F06); personality and behavioral disorders due to brain disease, damage, or dysfunction (F07); unspecified organic or symptomatic mental disorders (F09); disorders due to psychoactive substance use (F10-F19); schizophrenia, schizotypal, and delusional disorders (F20-29); manic episodes (F30); and bipolar affective disorder (F31). We also excluded individuals who had sought treatment for “nerves, anxiety, tension, or depression” from either their general practitioner or a psychiatrist. After exclusions, the UKBB dataset comprised 15,978 individuals aged 45 to 82 years, with a mean age of 64.26 ± 7.55 years and a median age of 65 years, including 7,244 females and 8,734 males (36,37). All UKBB participants provide informed consent as part of the ethical oversight maintained by a dedicated Ethics and Guidance Council, which collaborates with UKBB to uphold an Ethics and Governance Framework. Additionally, the study received IRB approval from the Northwest Multicenter Research Ethics Committee.

The HCP-YA dataset included 833 individuals aged 22 to 35 years, with a mean age of 28.66 ± 3.66 years and a median age of 29 years, consisting of 443 females and 390 males (39). The HCP-A dataset included 711 individuals aged 36 to 100 years, with a mean age of 60.54 ± 15.68 years and a median age of 58.67 years, consisting of 403 females and 308 males (40). Collectively, the datasets encompassed 17,522 individuals aged 22 to 100 years, with a mean age of 62.42 ± 10.96 years and a median age of 64 years, including 8,090 females and 9,432 males. All subject recruitment procedures and informed consent forms, including consent to share de-identified data, were approved by the Washington University IRB. Notably, the HCP datasets provided around 15 minutes of resting-state fMRI data per session, while the UKBB dataset included around 6 minutes of resting-state fMRI data. Because these data set contain multiple scans per participant, there are a total of 22,569 distinct resting-state fMRI scans in the training set. **Table 1** shows the demographic information of both training and test dataset.

**Table 1.** The Training Data set is composed of UKBB, HCP, and HCP-A datasets, while the FBIRN data set composes the Test Data.

|  |  | **N** | **Age (M ± SD)** | **Sex (M/F)** |
| --- | --- | --- | --- | --- |
| | All Training Data | 17522 | 62.42 ± 10.96 | 8090 / 9432 |
| **Training** | UKBB | 15978 | 64.26 ± 7.55 | 8734 / 7244 |
| | HCP | 833 | 28.66 ± 3.66 | 390 / 443 |
| | HCP-A | 711 | 60.54 ± 15.68 | 308 / 403 |
| **Test** | FBIRN | 311 | 37.88 ± 11.28 | 230 / 81 |

Additionally, data from the Functional Imaging Biomedical Informatics Research Network (FBIRN) were employed to estimate the brain age gap (BAG) and examine its link with cognitive metrics such as working memory and attention (41). For the FBIRN dataset, raw imaging data were collected across seven sites: University of California, Irvine; University of California, Los Angeles; University of California, San Francisco; Duke University/University of North Carolina at Chapel Hill; the University of Iowa; the University of Minnesota; and the University of New Mexico. Each participant provided written informed consent, with protocols approved by the institutional review boards at each respective site. All subjects with schizophrenia (SZ) were clinically stable at the time of scanning. Diagnoses were validated through the Structured Clinical Interview for DSM-IV (SCID-IV), while healthy controls (HC) were assessed using the SCID-I/NP to confirm the absence of schizophrenia. Exclusion criteria for HC included a current or history of major neurological or psychiatric disorders and having a first-degree relative with an Axis-I psychotic disorder, as determined by SCID evaluations. The FBIRN dataset contains individuals between 18 and 62 years old, with a mean age of 37.88 and a median age of 38. Out of 311 individuals in the set, 151 of them have a diagnosis of schizophrenia; these participants are between 18 and 62 years old, with a mean age of 38.77 and a median age of 39. Overall, there were 81 female participants and 230 male participants; among those diagnosed with schizophrenia, there were 36 female participants and 115 male participants. For the FBIRN dataset, written informed consent was obtained from all participants. Institutional review boards approved the consent process of each study site. **Table 1** presents the demographic information for all FBIRN participants, while **Supplementary Table 1** details the demographic and clinical data for each site separately. **Figure 1A** illustrates the count density of the training datasets (UKBB, HCP-YA, and HCP-A), while **Figure 1B** displays the count density of the test dataset (FBIRN) used in our study.

**Figure 1.**
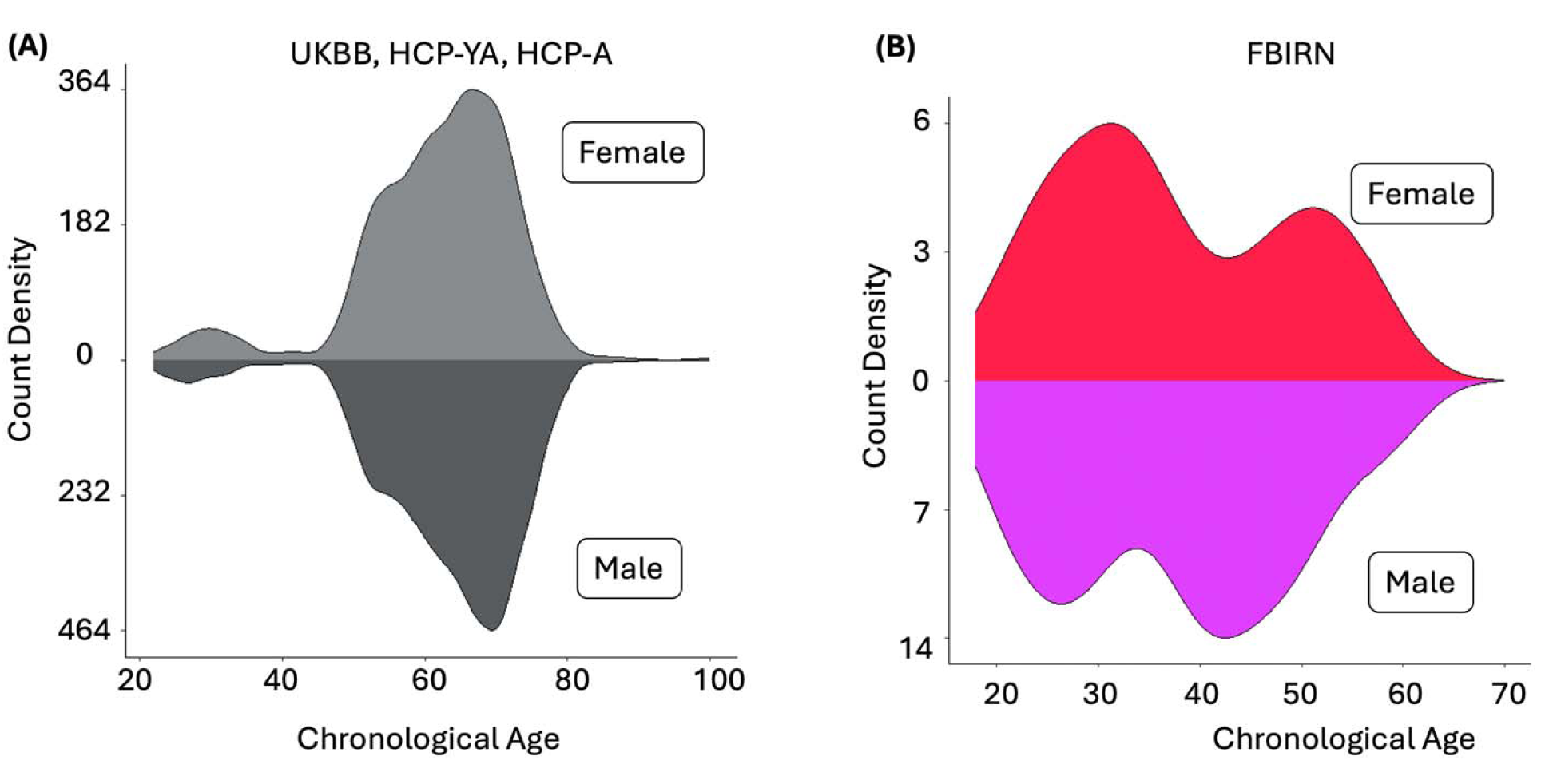
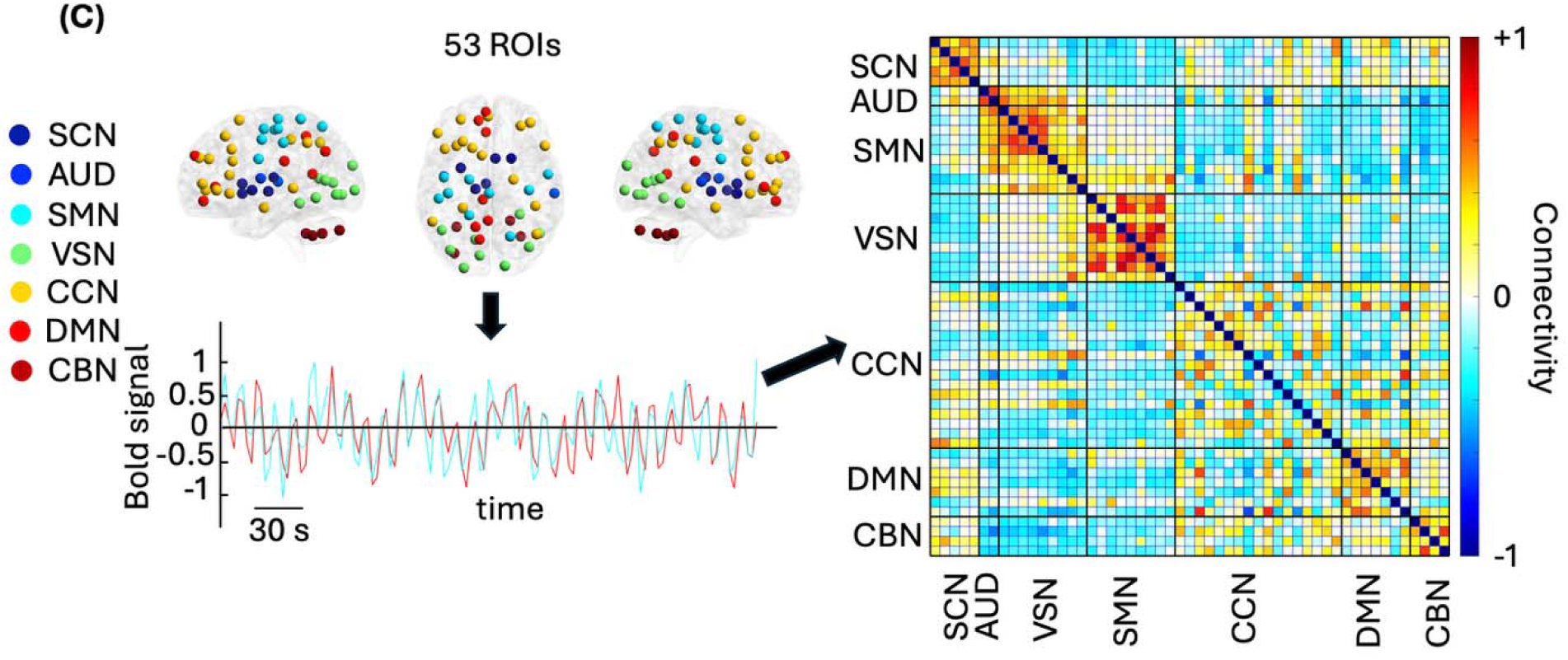
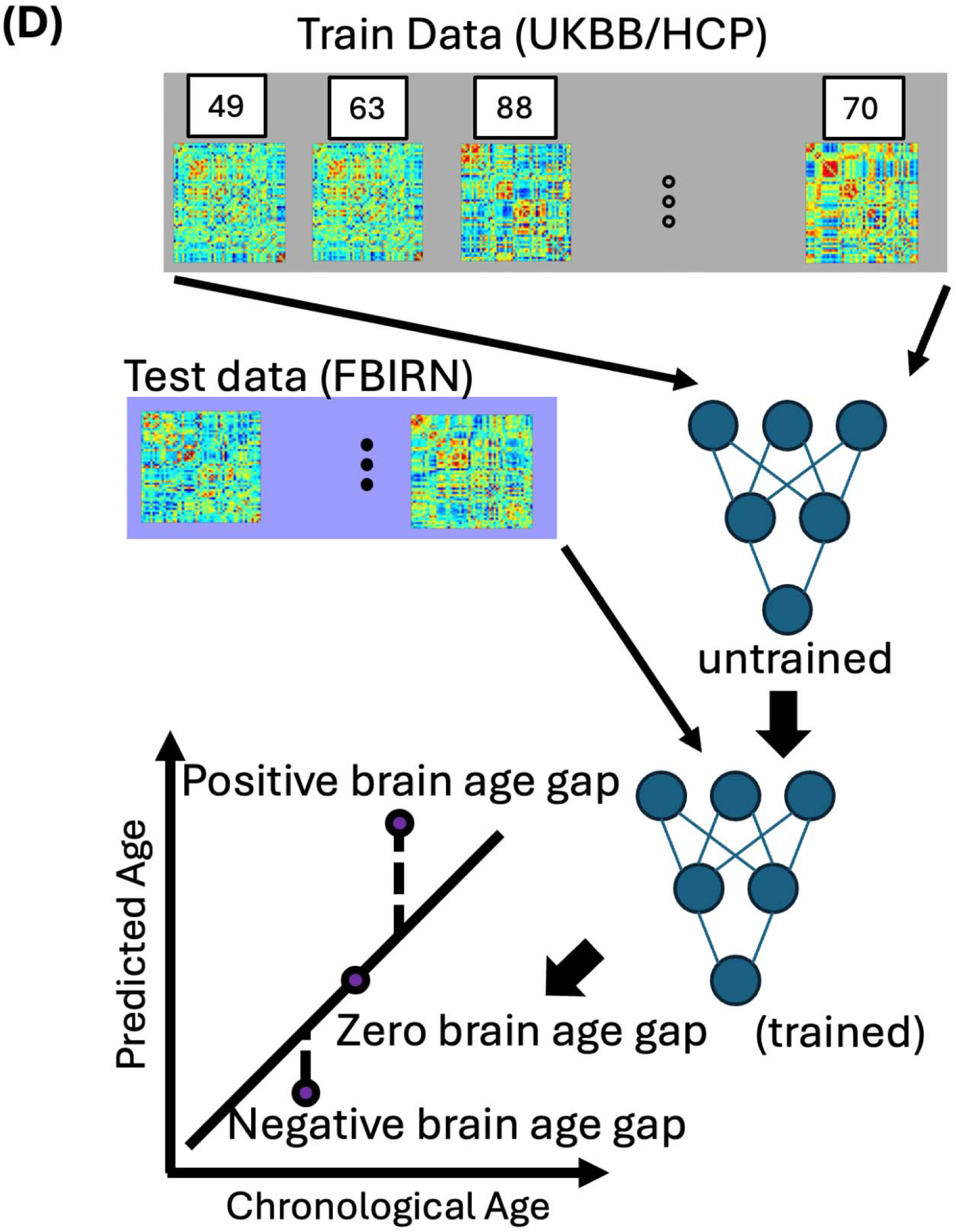
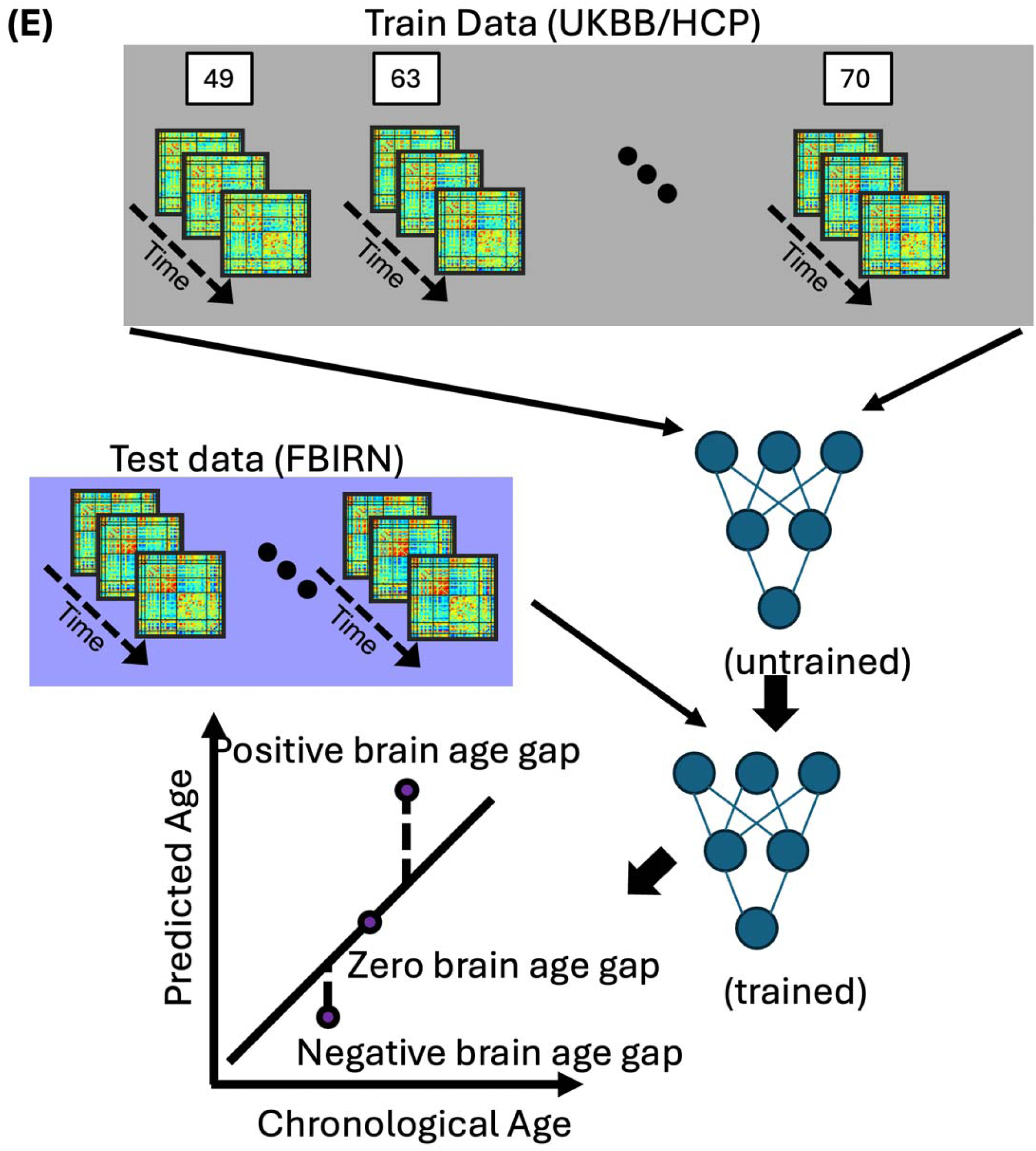
**A)** Count density of the training dataset used in the brain age prediction model employing both sFNC and dFNC variables. The training set comprises 17,522 individuals aged 22 to 100 years, including 8,090 females and 9,432 males. **B)** Count density of the FBIRN dataset used as test data to examine the link between BAG and cognitive/attention measures. The dataset includes 311 individuals aged 18 to 62 years (81 females and 230 males); our analysis focused on those with schizophrenia. **C)** NeuroMark pipeline extraction of 53 regions across seven networks: subcortical (SCN), auditory (AUD), sensorimotor (SMN), visual (VSN), cognitive control (CCN), default mode (DMN), and cerebellar (CBN). Both static and dynamic functional network connectivity (FNC) were computed between each pair of regions using the NeuroMark template, yielding a 53×53 connectivity matrix. A representative mean FNC was estimated from UK Biobank participants. **D)** Illustration of sFNC data processing: Training data (UKBB, HCP, and HCP-Aging) are input into an untrained model with chronological age as the label. The trained model then predicts brain age from test data (FBIRN). Comparison of predicted versus chronological age determines the brain age gap. **E)** Illustration of the dFNC data processing, analogous to panel D, demonstrating the training and application of the model using dynamic connectivity data.

### 2.2 Imaging Protocol

For UKBB, the imaging data were collected using Siemens Skyra 3T scanners. The resolution was 2.4 × 2.4 × 2.4 mm^3^. The data was collected as 490 timeframes over 6 minutes, with a TR of 0.735 s, a TE of 39ms, and a flip angle of 52°. During the scan, participants are instructed to fixate on a cross displayed on a screen to minimize eye movement and maximize data consistency. For HCP, the imaging data were collected using a Siemens 3T scanner as 1200 frames per run over 14:33 minutes per run, with 4 runs total. The TR was 720 ms, the TE was 33.1, the flip angle was 52°, and the slice thickness was 2.0mm.

For the HCP-YA, imaging data were captured using a 3T Siemens Prisma scanner equipped with a 32-channel phased-array head coil. The rs-fMRI acquisition parameters included a TR of 720 ms, a TE of 33.1 ms, and a field of view of 208 × 180 mm². A flip angle of 52° was used, with images obtained across 72 oblique-axial slices at a resolution of 2.0 × 2.0 × 2.0 mm³. Each scanning session was conducted over a duration of approximately 14 minutes and 40 seconds. For the HCP-A imaging data were collected using a 3T Siemens Prisma scanner equipped with a 32-channel phased-array head coil. The rs-fMRI protocol included the following parameters: TR of 800 ms, TE of 37 ms, a field of view of 810 × 936 mm², and a flip angle of 52°. Images were acquired with a resolution of 2.0 × 2.0 × 2.0 mm³ across 72 oblique-axial slices. Each scanning session lasted approximately 14 minutes and 40 seconds. Similar to the UKBB, participants in the HCP are asked to fixate on a cross presented on a screen during the scan to reduce eye movements and ensure consistent data collection.

For FBIRN, imaging data were acquired using six Siemens 3T scanners and one General Electric 3T scanner. All sites followed a uniform rs-fMRI protocol. T2*-weighted functional images were captured using an echo-planar imaging sequence, aligned along the anterior and posterior commissure (AC-PC) line, with the following parameters: TE of 30 ms, TR of 2 s, flip angle of 77°, slice gap of 1 mm, voxel dimensions of 3.4 × 3.4 × 4 mm³, and a series of 162 frames spanning 5 minutes and 38 seconds. During scans, participants were instructed to keep their eyes closed.

### 2.3 Data Processing

Data from fMRI were preprocessed using Statistical Parametric Mapping (SPM12, https://www.fil.ion.ucl.ac.uk/spm/) in the MATLAB 2019 environment. We conducted a rigid body motion correction with SPM’s toolbox to address head motion. Subsequently, the imaging data were spatially normalized to an echo-planar imaging (EPI) template in standard Montreal Neurological Institute (MNI) space. Finally, a Gaussian kernel with a full width at half maximum (FWHM) of 6 mm was applied to smooth the fMRI images.

### 2.4 Extracting independent components using NeuroMark

To obtain reliable independent components (ICs), we employed the NeuroMark fully automated ICA pipeline, which integrates previously derived component maps as spatial constraints. The NeuroMark framework utilizes templates developed from substantial datasets, specifically the Human Connectome Project (HCP: https://www.humanconnectome.org/study/hcp-young-adult/document/1200-subjects-data-release, 823 subjects after the subject selection) and the Genomics Superstruct Project (GSP: https://dataverse.harvard.edu/dataverse/GSP, 1005 subjects post-selection). This approach has proven effective across numerous studies, identifying a broad array of imaging markers for various brain disorders. Further information on template development is available in our prior publication on the NeuroMark method (42). The NeuroMark template includes 53 independent components (ICs), categorized into seven functional networks: subcortical (SCN), auditory (AUD), sensorimotor (SMN), visual (VSN), cognitive control (CCN), default-mode (DMN), and cerebellar networks (CBN), as shown in **Figure 1C** and **Supplementary Figure 1**. **Supplementary Table 2** also shows all 53 ICs and their interactions. For the ICA analysis, we utilized these templates via the NeuroMark_fMRI_1.0 template, accessible through GIFT v4.0.5.14 GIFT (http://trendscenter.org/software/gift and on the TReNDS website @ http://trendscenter.org/data). Additional denoising and artifact removal steps prior to calculating dynamic functional connectivity included: 1) linear, quadratic, and cubic de-trending; 2) multiple regression of the six realignment parameters and their temporal derivatives; 3) outlier removal; and 4) low pass filtering below a frequency of 0.15 Hz.

### 2.6 Static functional network connectivity

To estimate sFNC, we calculated the Pearson correlation between pairs of ICs in each subject as shown in equation 1

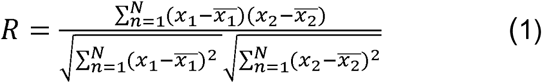

where *x*1 and *x*2 are time course signals and 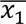 and 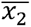 are the mean of *x*1 and *x*2, respectively. This Pearson correlation takes values in the interval [− 1, 1] and measures the strength of the linear relationship between *x*1 and *x*2. Each FNC is a 53×53 matrix, from which we derived a total of 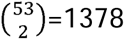 connectivity features. **Figure 1C** shows a representative FNC we used in our study.

### 2.7 Dynamic functional network connectivity

To compute dynamic functional network connectivity (dFNC), we employed a sliding window approach. We used a tapered window, formed by convolving a rectangular window with the duration of 20TR (UKBB: 14.7 s, HCP-YA: 14.4 s, HCP-A: 16 s, FBIRN: 40s) with a Gaussian kernel (σ = 3), to precisely focus on data at each time point (see **Supplementary Figure 2**) (30,31,33,34,48,50,52). This approach, detailed in Equation 1, was utilized to compute the FNC at each time point. We then aggregated the dFNC estimates for each window and each subject to construct a three-dimensional array (C × C × T), where C represents the 53 independent components, and T represents the number of windows. This array captures the temporal variations in connectivity among the independent components.

### 2.8 Training the model

In our study, the resting-state fMRI time series were uniformly downsampled to the shortest duration across datasets, corresponding to approximately 5.4 minutes. We then computed dFNC matrices using a sliding window approach with a window size of 40 TRs. To evaluate model stability, K=5-fold cross-validation was performed on the training set of 22,569 scans from the UKB, HCP-YA and HCP-A data sets. For sFNC, we deployed a brain connectivity graph convolutional network or BCGCN to predict the brain age (43,44). The BCGCN model utilized ReLU activation for all layers except the output, which employed a linear activation (see **Supplementary Figure 3A**). For the analysis of the dFNC data, we employed a bi-directional Long Short-Term Memory network (biLSTM) (45) configured with three recurrent layers with 128 hidden units each, a dropout rate of 0.1, and a fully connected regression layer to predict brain age (see **Supplementary Figure 3B**). In both models, we utilized an Adam optimizer targeting Mean Absolute Error, with a learning rate of 1e^-3^ and a batch size of 64, conducting training over 100 epochs. The epoch with the optimal cross-validation performance was selected for inference on the test set. **Figure 1D** and **Figure 1E** illustrate the modeling and testing procedures for the sFNC and dFNC features, respectively.

### 2.9 Statistical analysis

We developed brain age prediction models incorporating both sFNC and dFNC variables. These models were then applied to the FBIRN dataset to estimate the predicted brain age and subsequently brain age gap (BAG) for each participant. To investigate the associations between these BAGs and cognitive measures, including working memory and attention vigilance, we constructed a General Linear Model (GLM). This model included age, sex, site, age^2^, the interaction of age and sex, and the diagnosis (schizophrenia or control) as covariates. For each cognitive measure, we tested two hypotheses—assessing the association of BAG derived from two modalities (sFNC-based and dFNC-based BAGs). We corrected for multiple comparisons across the two modalities using the False Discovery Rate (FDR) correction method to ensure the robustness of our findings (46).

## 3. RESULTS

### 3.1 Training and validating the brain age prediction model on the healthy population

After developing brain age prediction models using wide-brain static and dynamic functional network connectivity (sFNC and dFNC, respectively), we validated their performance on a validation dataset within each run. This step was essential to assess the models’ generalizability and accuracy in predicting brain age. We calculated the Pearson correlation coefficients between the chronological brain age and the predicted brain age to evaluate the models’ performance. Results demonstrated a strong correlation between the predicted brain age from the sFNC-based model and chronological age, with a correlation value of 0.8755 (**Figure 2A**). Similarly, the dFNC-based model also showed a strong correlation, with a correlation value of 0.8675, illustrating its effectiveness in age prediction (**Figure 2B**).

**Figure 2.**
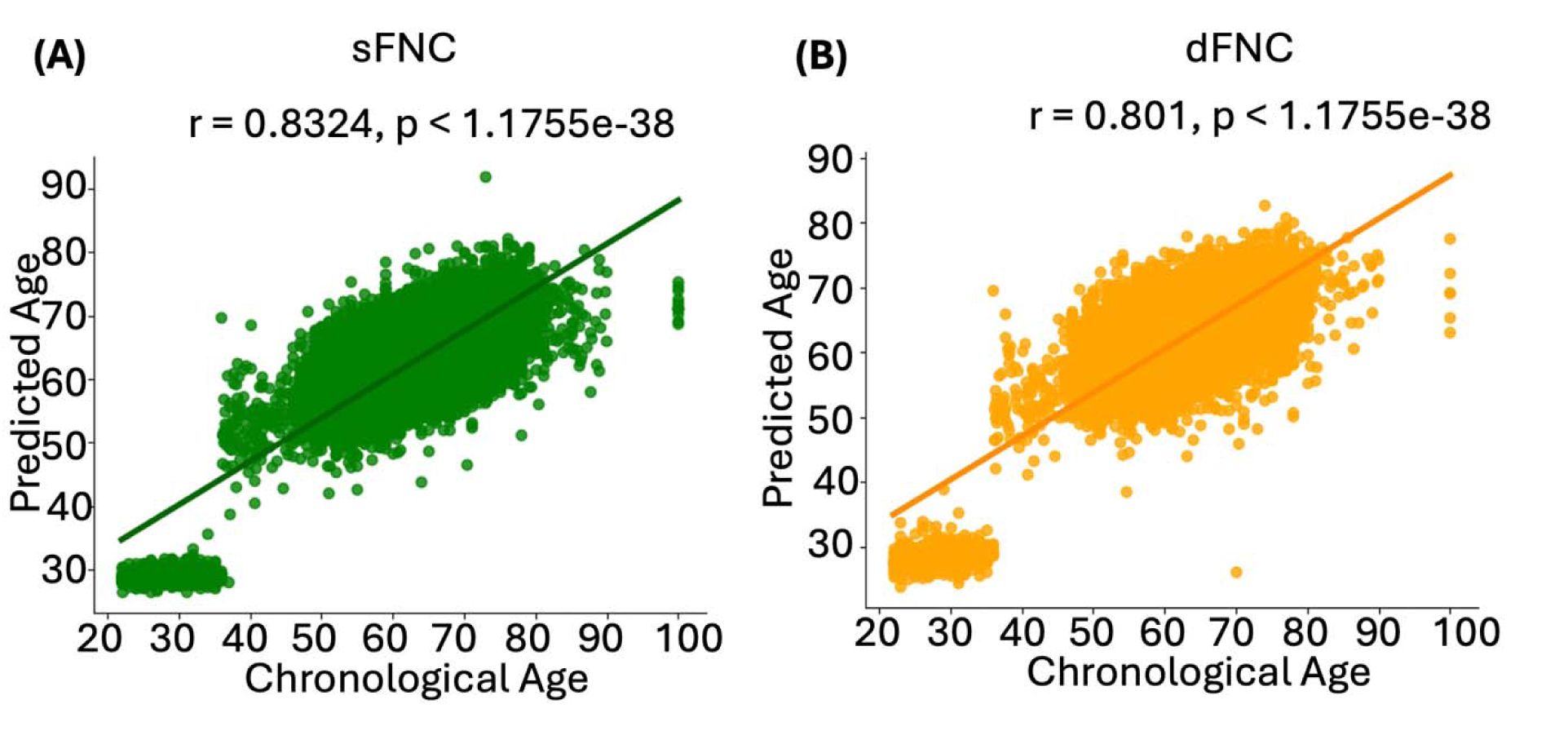
Correlation between brain age predictions and chronological age in sFNC- and dFNC-based models. These graphs illustrate the relationship between chronological age and predicted brain age in the training dataset (UK Biobank, HCP, and HCP Aging). **A)** sFNC-based predictions (green) show a strong positive correlation with chronological age (r = 0.8324, p < 1.1755e^-38^). **B)** dFNC-based predictions (orange) also show a significant positive correlation (r = 0.801, p < 1.1755e^-38^).

### 3.2 Static and dynamic FNC-based wide-brain age gap links with attention vigilance

After developing and validating the brain age prediction models, we applied them to the FBIRN dataset to calculate wide-brain BAGs (wBAGs), representing brain age gaps estimated from wide-brain sFNC and dFNC for each individual. We then constructed GLMs to examine the association between wBAGs and attention vigilance while adjusting for age, sex, study site, age^2^, the age-by-sex interaction, and diagnosis as covariates. The corresponding p-values were further adjusted to account for testing two modalities. Because our age prediction model performs less accurately for individuals under 38 years old (see **Figure 2**), we restricted the FBIRN analysis to participants over 38. This resulted in a final sample of 153 participants (121 males) with a mean age of 47.73□±5.94 years. **Figure 3A** shows a significant negative association between sFNC-based wBAGs and attention vigilance (*r*□=-0.2923, *β*□= −0.9860, *SE*=0.2829, *95% CI*: −1.5454 to −0.4263, *FDR p*=0.0013, *N*=153). **Figure 3B** demonstrates a significant association between dFNC-based wBAGs and attention vigilance (*r*□=□-0.2715, *β*=-0.4395, *S*□=0.1366, *95% CI*: −0.7099 to −0.1691, *FDR p*=0.0016, *N*□=□153). In both models, we observed a negative association between wBAGs and cognitive performance, indicating that individuals with older-appearing brains tend to perform worse on attention vigilance tasks.

**Figure 3.**
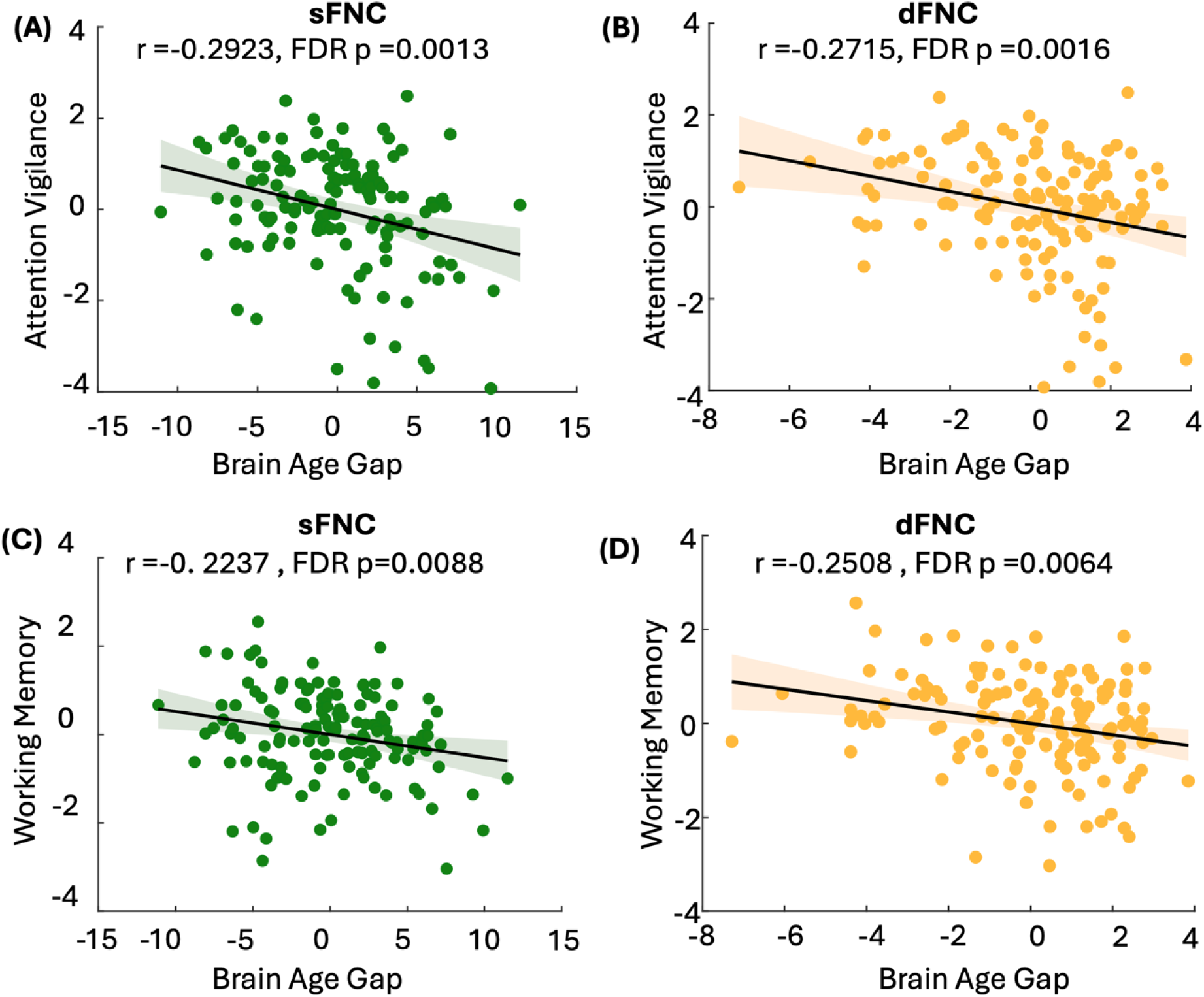
Associations between sFNC- and dFNC-based wide-brain age gap (wBAG) and cognitive performance in individuals with schizophrenia. After developing and validating the brain age prediction models, we applied them to the FBIRN dataset to calculate wBAGs based on static (sFNC) and dynamic (dFNC) functional network connectivity features. Analyses were restricted to individuals older than 38 years due to reduced model accuracy in younger participants (see **Figure 2**), resulting in a final sample of 160 participants (mean age = 47.73□±□5.94 years; 121 males). General Linear Models (GLMs) were used to examine associations between wBAGs and two cognitive domains—attention vigilance and working memory—controlling for age, sex, study site, age squared, age-by-sex interaction, and diagnosis. P-values were adjusted for multiple comparisons across modalities using the Benjamini-Hochberg FDR correction. **A)** sFNC-based wBAG was significantly negatively associated with attention vigilance (*r*=–0.2923, *β*=–0.9860, *SE*=0.2829, *95% CI*: –1.5454 to – 0.4263, *FDR p*=0.0013, *N*=153). **B)** dFNC-based wBAG was significantly negatively associated with attention vigilance (*r*=–0.2715, *β*=–0.4395, *SE*=0.1366, *95% CI*: –0.7099 to –0.1691, *FDR p*=0.0016, N=153). **C)** sFNC-based wBAG was significantly negatively associated with working memory (*r*=–0.2237, *β*=–0.9603, *SE*=0.3615, *95% CI*: –1.6761 to –0.2445, *FDR p*=0.0088, *N*=153). D) dFNC-based wBAG was significantly negatively associated with working memory (*r*=–0.2508, *β*=–0.5195, *SE*=0.1732, *95% CI*: –0.8621 to –0.1769, *FDR p*=0.0064, *N*=153). Across both cognitive domains, higher wBAGs—reflecting accelerated brain aging—were associated with poorer cognitive performance.

### 3.3 Static and dynamic FNC-based wide-brain age gap links with working memory

We constructed GLMs to examine the association between sFNC- and dFNC-based wBAGs and working memory while adjusting for age, sex, study site, age^2^, age-by-sex interaction, and diagnosis as covariates. P-values were further corrected for multiple comparisons across the two modalities. **Figure 3C** shows a significant negative association between sFNC-based wBAGs and working memory (*r*=-0.2237, *β*=-0.9603, *SE*=0.3615, *95% CI*=-0.2918 to −0.0856, *FDR p*=0.0088, *N*=153). **Figure 3D** demonstrates a significant negative association between dFNC-based wBAGs and working memory (*r*=-0.2508, *β*=-0.5195, *SE*=0.1732, *95% CI*= −0.8621 to −0.1769, *FDR p*=0.0064, *N*=151). These negative correlations indicate that individuals with higher brain age relative to chronological age tend to exhibit poorer working memory performance.

### 3.4 Sub-network brain age gaps predict attention vigilance and working memory

The next question was whether sFNC- and dFNC-based sub-network brain age gaps or subBAGs predict attention vigilance, and working memory performance. To address this, we developed brain age prediction models separately for each network using data from our healthy population. Specifically, we created seven distinct brain age prediction models corresponding to seven networks including SCN, AUC, SMN, VSN, CCN, DMN, and CBN. The correlation between chronological age and predicted brain age for each sub-network is presented in **Supplementary Figure 4**. Next, we examined the association between subBAGs and attention vigilance, and working memory, controlling for age, sex, site, age², age-by-sex interaction, and diagnosis as covariates. Given seven networks and two modalities (sFNC and dFNC), a total of 14 statistical tests were performed, and p-values were corrected accordingly.

**Figure 4.A** illustrates the relationship between sFNC- and dFNC subBAGs and attention vigilance. Each bar represents the association between subBAGs and attention vigilance, with crosshatched bars indicating dFNC-based subBAGs and solid bars representing sFNC-based subBAGs. Bars showing a significant association after FDR correction are marked with double asterisks. As shown, the sFNC-based subBAGs estimated from the subcortical network exhibit a negative association with attention vigilance (*r* = −0.2357, *β* = −0.1813, *SE* = 0.0655, *95% CI*: - 0.3111 to −0.0516, *FDR p* = 0.0228, *N* = 153). Similarly, the dFNC-based subBAGs from the subcortical network show a significant negative association with attention vigilance (*r*=-0.2806, *β*=-0.2989, *SE*=0.0896, *95% CI*: −0.4763 to −0.1215, *FDR p*=0.0140, *N*=153). Additionally, dFNC-based subBAGs estimated from the sensorimotor network show a significant negative association (*r* = −0.2384, *β*=-0.2153, *SE*=0.0769, *95% CI*: −0.3675 to −0.0631, *FDR p*=0.0228, *N*=153) and from the default mode network (*r* =-0.2666, *β* =-0.1053, *SE*=0.0333, *95% CI*: - 0.1713 to −0.0392, *FDR p* =0.0140, *N*=153). Our results also indicate a significant negative association between sFNC-based subBAGs estimated from the cognitive control network and attention vigilance (*r*=-0.2081, *β*=-0.0353, *SE*=0.0145, *95% CI*: −0.0641 to −0.0065, *FDR p*=0.0455, *N*=153). **Supplementary Table 3** presents the complete set of models examining the association between subBAGs and attention vigilance.

**Figure 4.**
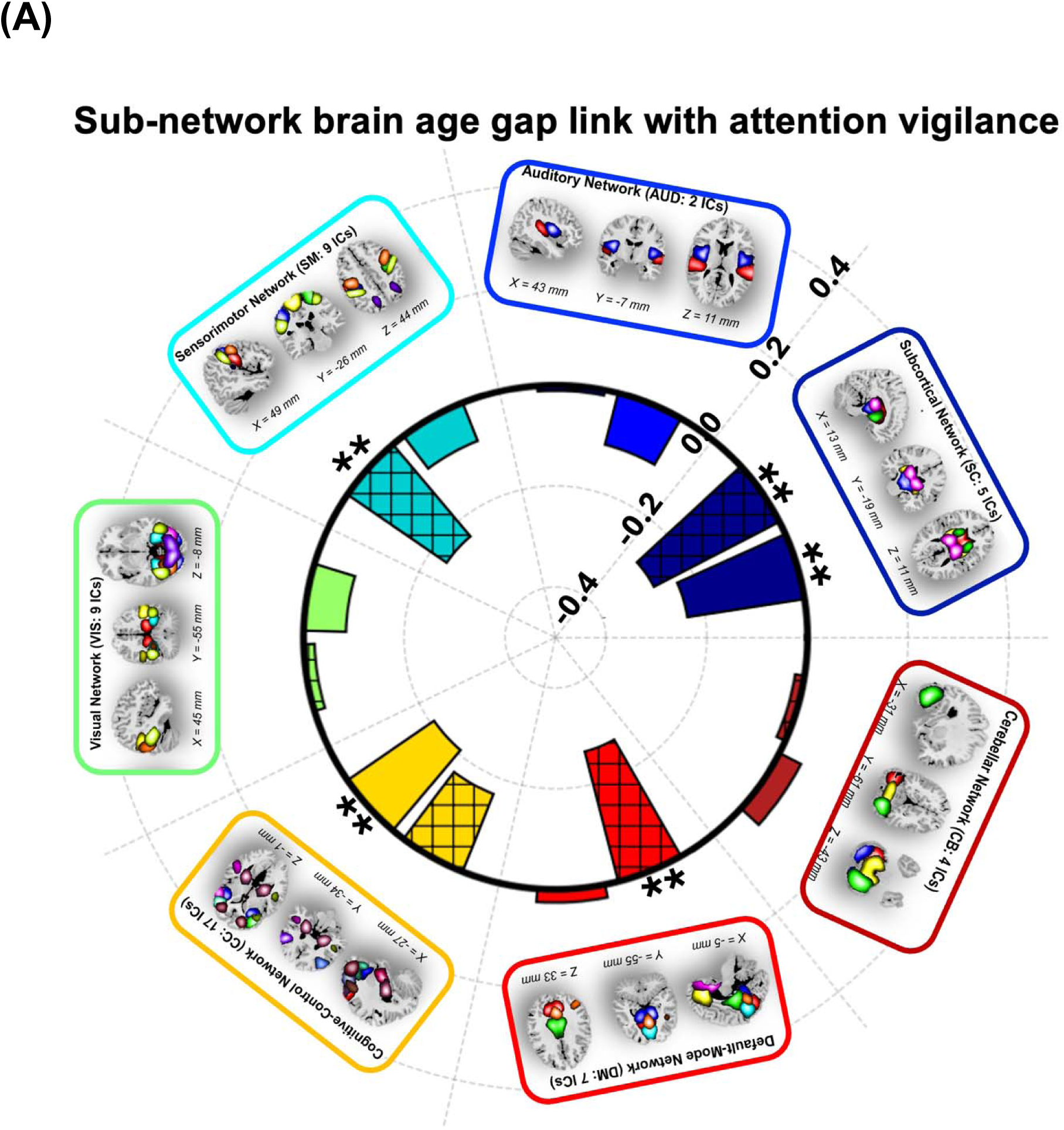
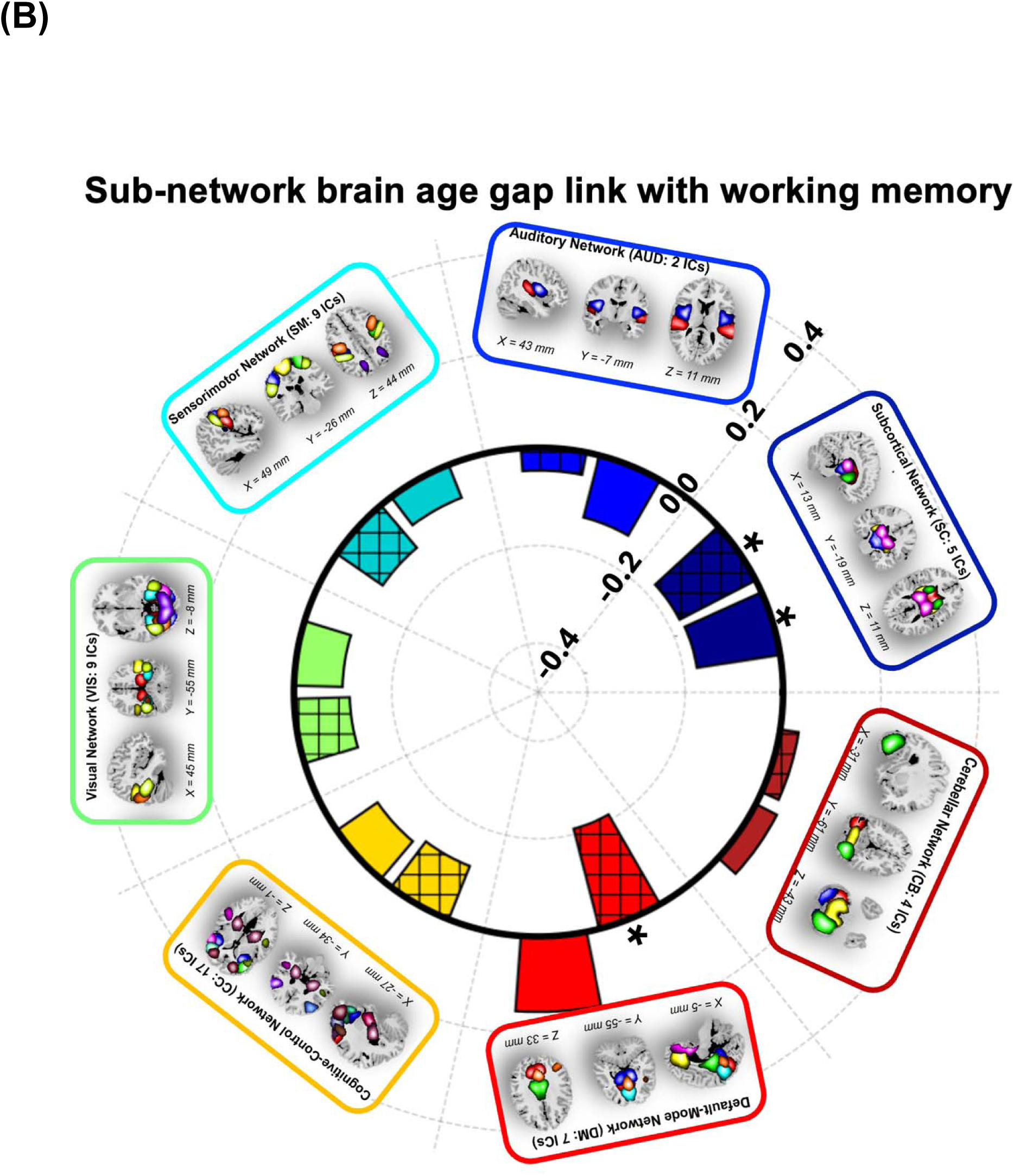
Associations between subBAGs and cognitive performance. **A)** Relationship between subBAGs and attention vigilance. Crosshatched bars represent *dFNC*-based subBAGs and solid bars represent sFNC*-*based subBAGs. Bars with double asterisks indicate significant associations after FDR correction. Significant negative associations were observed for *sFNC*- based subBAGs from the subcortical network (*r*=-0.2357, *β*=-0.1813, *SE*=0.0655, *95% CI*=-0.3111 to −0.0516, *FDR p*=0.0228, *N*=153) and dFNC*-*based subBAGs from the subcortical network (*r*=-0.2806, *β*=-0.2989, *SE*=0.0896, 95% *CI*=-0.4763 to −0.1215, *FDR p*=0.0140, *N*=153). Additional significant negative associations were observed for *dFNC*-based subBAGs from the sensorimotor network (*r*=-0.2384, *β*=-0.2153, *SE*=0.0769, 95% *CI*=-0.3675 to −0.0631, *FDR p*=0.0228, *N*=153) and the default mode network (*r*=-0.2666, *β*=-0.1053, *SE*=0.0333, 95% *CI*=-0.1713 to −0.0392, *FDR p*=0.0140, *N*=153). The sFNC-based subBAGs from the cognitive control network also showed a significant negative association (*r*=-0.2081, *β*=-0.0353, *SE*=0.0145, 95% *CI*=-0.0641 to −0.0065, *FDR p*=0.0455, *N*=153). **B)** Relationship between subBAGs and working memory. Significant negative associations (*uncorrected p*<0.05) were identified for sFNC*-*based subBAGs from the subcortical network (*r*=-0.1724, *β*=-0.1050, *SE*=0.0518, 95% *CI*=-0.2076 to −0.0020, *uncorrected p*=0.0447, *N*=153), *dFNC*-based subBAGs from the subcortical network (*r*=-0.1754, *β*=-0.1473, *SE*=0.0714, 95% *CI*=-0.2885 to −0.0060, *uncorrected p*=0.0410, *N*=153), and dFNC-based subBAGs from the default mode network (*r*=-0.2114, *β*=-0.0662, *SE*=0.0264, 95% *CI*=-0.1186 to −0.0139, *uncorrected p*=0.0134, *N*=153). However, none of these associations survived FDR correction. All models were adjusted for age, sex, site, age², age-by-sex interaction, and diagnosis.

**Figure 4.B** shows the link between sFNC- and dFNC-based subBAGs with working memory using GLM with age, sex, site, age², age-by-sex interaction, and diagnosis as covariates. Among all models, we identified only three showing a significant association between subBAGs and working memory (as shown with single asterisk): sFNC-based subBAGs estimated from the subcortical network (*r*=-0.1724, *β* = −0.1050, *SE* = 0.0518, *95% CI*: −0.2076 to −0.0020, *uncorrected p*=0.0447, *N*=153), dFNC-based subBAGs estimated from the subcortical network (*r* =-0.1754, *β*=-0.1473, *SE*=0.0714, *95% CI*: −0.2885 to −0.0060, *uncorrected p*=0.0410, *N*=153), and dFNC-based subBAGs estimated from the default mode network (*r* =-0.2114, *β* =-0.0662, *SE*=0.0264, *95% CI*: −0.1186 to −0.0139, *uncorrected p*=0.0134, *N*=153). However, none of these associations remained significant after FDR correction. **Supplementary Table 4** presents the complete set of models examining the association between subBAGs and working memory.

## DISCUSSION

The brain-age gapD(BAG), the difference between a person’s predicted brain age and chronological age, has emerged as a compact indicator of neurobiological health: a larger positive BAG signals accelerated ageing, whereas a negative BAG reflects resilience.

Schizophrenia reliably shows an elevated BAG, and larger BAGs correlate with poorer working-memory performance□ (47,49,51). Crucially, almost all BAG work to date relies on structural MRI or static FNC, overlooking the millisecond-scale reconfiguration of brain networks. We present the first dynamic-FNC brain age models—at both wide-brain (wBAG) and network-specific (subBAG) levels—leveraging time-resolved connectivity to capture the neural volatility that likely underlies cognitive dysfunction. By testing dFNC-BAG against working memory and attention vigilance performance in schizophrenia cohort, we aim to uncover a previously inaccessible layer of pathophysiology and position dFNC-BAG as new precision biomarker capable of guiding circuit-based interventions and accelerating translational psychiatry.

Extant research shows that sustained attention declines with normal aging (53), and schizophrenia magnifies this decline, with patients repeatedly scoring well below normative thresholds across sites, ages, and sexes□ (54). Our data reveal a robust negative association between attention vigilance measure and both wBAG and subBAG indices of brain age acceleration—whether derived from static or dynamic FNC. Put simply, the “older” the brain looks, the poorer the patient performs. This powerful link spotlights accelerated macro- and micro-scale brain aging as a driving force behind attentional breakdown in schizophrenia and elevates BAG to a precision and network-level biomarker with clear therapeutic implications. Convergent neuroimaging work has already mapped attentional control to frontoparietal and default mode circuitry; our findings now tie those circuits’ temporal dysmaturation directly to real-world cognitive deficits. Neuroimaging studies have localized specific brain regions and connectivity networks that facilitate attentional processes. For instance, processing temporal stimuli has been linked to activity in the pre-supplementary motor area (55), whereas joint attention engages the ventromedial frontal cortex, cingulate cortex, caudate nuclei, and left superior frontal gyrus (56). Mirroring these maps, our strongest BAG-attention associations localize to sensorimotor and subcortical networks—notably the caudate, thalamus, and putamen—underscoring their central role in the disorder’s cognitive burden. Overall, this work bridges neuroanatomical change with clinical symptomatology and supports BAG as a precision tool for early identification and stratification in neuropsychiatric disorders.

Working memory deficits—spanning visuospatial, phonological, and executive domains—are a core cognitive feature of schizophrenia, consistently observed across cohorts relative to healthy controls (57). These deficits are compounded by normative age-related decline, with working memory performance deteriorating progressively across adulthood (58). Building on this foundation, we examined the relationship between BAG and working memory using both sFNC and dFNC models. Both sFNC- and dFNC-based wBAG showed robust negative associations with working memory performance in the FBIRN cohort. In other words, greater brain age acceleration predicted worse working memory capacity. This finding extends prior work by linking functional BAG to cognition in schizophrenia, reinforcing the relevance of BAG as a mechanistic marker of cognitive vulnerability. Notably, our results highlight the subcortical and default mode networks as key contributors to this relationship, pointing to specific neural systems where accelerated ageing most profoundly impacts working memory function.

Notably, the association between wBAGs and working memory, and subBAGs and attention vigilance, was stronger for dFNC-derived BAG, suggesting cognitive deficits in schizophrenia may be more closely linked to dynamic reconfiguration of brain networks than to static connectivity patterns. This aligns with evidence that cognition depends on flexible, moment-to-moment network adaptations to meet cognitive demands (59). By capturing temporal fluctuations in connectivity, the dFNC approach provides a richer, more ecologically valid representation of brain function, making it more sensitive to the cognitive disruptions observed in schizophrenia. In contrast, sFNC averages connectivity over time, potentially obscuring critical dynamic alterations. The higher temporal resolution of dFNC also yields more data points per scan, enhancing the detection of subtle brain–behavior associations (30,31,33). These findings highlight the unique potential of dFNC-based BAG as a precision biomarker to capture clinically meaningful cognitive decline and inform individualized therapeutic strategies in schizophrenia.

Our study has several limitations that merit consideration. Firstly, the choice of window size in dynamic connectivity analysis inherently assumes certain characteristics about temporal dynamics. Shorter windows capture rapid fluctuations effectively, while longer windows tend to smooth these fluctuations, potentially obscuring meaningful variability. Future research should explore a broader range of window sizes to more comprehensively assess their impact on brain connectivity measures (60). Additionally, our study excluded participants with any health conditions from the UKBB dataset. Future studies may benefit from including a broader cohort by aggregating data from multiple datasets that include healthy individuals, which could enhance the generalizability and robustness of the findings. Moreover, prior research suggests that factors like increased income or engagement in cognitively stimulating activities can mitigate age-related declines in working memory. However, our model did not account for variables such as average activity levels, diet, or other environmental factors that could also influence cognitive and brain function(62–64). Adjusting both brain and cognitive measures for these variables in the future studies could enhance our understanding of the complex interactions between lifestyle, health conditions, and cognitive aging, providing a more nuanced perspective on these relationships. Finally, due to deviations observed in our brain-age predictions for younger adults, likely driven by the limited representation of this age group in our healthy training dataset, we excluded FBIRN participants under 38. Expanding brain-age modeling efforts to include more younger adults from diverse datasets is an important next step, enabling exploration of whether BAG meaningfully captures cognitive deficits earlier in life and across the full adult lifespan.

In conclusion, our study establishes BAG, derived from both static and dynamic functional connectivity at wide-brain and subnetwork levels, as a powerful and clinically relevant marker of cognitive deficits in schizophrenia. We demonstrate that higher BAG—reflecting accelerated brain aging—is significantly associated with worse working memory and attention vigilance, capturing core cognitive dysfunctions of the disorder. Importantly, the dynamic approach provides unique sensitivity by incorporating temporal fluctuations in connectivity, offering deeper insight into the brain’s functional organization than traditional static methods. Our findings highlight dFNC-based BAG as especially informative, uncovering stronger links to the neural substrates of cognitive decline. These results not only advance mechanistic understanding but also open new avenues for clinical translation. Future research expanding dynamic modeling and integrating lifestyle and health factors holds tremendous promise for optimizing prediction models. Overall, this work lays the foundation for next-generation precision psychiatry tools capable of early detection and personalized intervention strategies to mitigate cognitive deterioration and improve long-term outcomes in schizophrenia.

## Data Availability

All data produced in the present study are available upon reasonable request to the authors

## Code and data availability statement

The code used for preprocessing and FNC calculation are available at https://trendscenter.org/software/. Also, statistical parametric mapping (SPM 12) is available at https://www.fil.ion.ucl.ac.uk/spm/. The Neuromark framework and the Neuromark template (Neuromark_fMRI_1.0) have been made available and incorporated into the Group ICA Toolbox (https://github.com/trendscenter/gift). Users worldwide can now directly download and utilize these resources. We also use this https://www.nitrc.org/projects/bnv/ for brain graph.

## Acknowledgements

We thank the participants of this study and those involved in data collection. This work was supported by the National Institutes of Health (NIH) grants R01MH123610 and T32MH125786, the National Science Foundation (NSF) grant 2112455, and the Phyllis and Jerome Lyle Rappaport Mental Health Research Scholars Award.

## Competing interests

Dr. Sendi has served as a consultant for Niji Corp for unrelated work. Dr. Mathalon has served as a consultant for Aptinyx, Boehringer-Ingelheim Pharmaceuticals, Cadent Therapeutics, and Greenwich Biosciences for unrelated work. The remaining authors declare no competing interests.

## Notes

### Author Declarations

Ethics committee/IRB of the Northwest Multicentre Research Ethics Committee (UK Biobank), the Washington University Institutional Review Board (Human Connectome Project), and the Institutional Review Boards at University of California Irvine, University of California Los Angeles, University of California San Francisco, Duke University/University of North Carolina at Chapel Hill, University of Iowa, University of Minnesota, and University of New Mexico (FBIRN) gave ethical approval for this work.

