## Supplementary Information for "Functional Brain Age Acceleration from Dynamic and Static Connectivity Predicts Working Memory and Attention Deficits in Schizophrenia"

|  |  | **SZ** | **HC** | **P-value** |
| --- | --- | --- | --- | --- |
| **FBRIN**  **Site 1** | **Number** | 21 | 28 | NA |
|  | **Age** | 30.04±8.60 | 34.78±9.40 | 0.1 |
|  | **Gender(M/F)** | 17/4 | 21/7 | 0.99 |
|  | **PANSS (positive)** | 15.72±5.57 | NA | NA |
|  | **PANSS(negative)** | 14.11±3.14 | NA | NA |
|  | **Attention vigilance** |  |  |  |
|  | **Working memory** |  |  |  |
| **FBIRN**  **Site 2** | **Number** | 12 | 10 | NA |
|  | **Age** | 44.91±11.34 | 38.10±9.39 | 0.14 |
|  | **Sex (M/F)** | 12/0 | 7/3 | 0.62 |
|  | **PANSS (positive)** | 16.90±6.70 | NA | NA |
|  | **PANSS negative)** | 17.20±7.39 | NA | NA |
|  | **Attention vigilance** |  |  |  |
|  | **Working memory** |  |  |  |
| **FBIRN**  **Site 3** | **Number** | 24 | 27 | NA |
|  | **Age** | 44.41±11.90 | 42.48±12.56 | 0.57 |
|  | **Sex (M/F)** | 19/5 | 21/6 | 0.99 |
|  | **PANSS (positive)** | 16.95±4.43 | NA | NA |
|  | **PANSS negative)** | 16.87±5.91 | NA | NA |
|  | **Attention vigilance** |  |  |  |
|  | **Working memory** |  |  |  |
| **FBIRN**  **Site 4** | **Number** | 26 | 26 | NA |
|  | **Age** | 36.88±12.82 | 35.23±10.56 | 0.61 |
|  | **Sex (M/F)** | 21/5 | 20/6 | 0.99 |
|  | **PANSS (positive)** | 14.29±3.77 | NA | NA |
|  | **PANSS (negative)** | 13.41±4.64 | NA | NA |
|  | **Attention vigilance** |  |  |  |
|  | **Working memory** |  |  |  |
| **FBIRN**  **Site 5** | **Number** | 14 | 15 | NA |
|  | **Age** | 36.64±10.27 | 37.53±9.76 | 0.81 |
|  | **Sex (M/F)** | 9/5 | 10/5 | 0.99 |
|  | **PANSS (positive)** | 13.14±4.46 | NA | NA |
|  | **PANSS (negative)** | 15±6.28 | NA | NA |
|  | **Attention vigilance** |  |  |  |
|  | **Working memory** |  |  |  |
| **FBIRN**  **Site 6** | **Number** | 29 | 27 | NA |
|  | **Age** | 36.27±11.08 | 34.51±10.85 | 0.54 |
|  | **Gender(M/F)** | 17/12 | 16/9 | 0.99 |
|  | **PANSS (positive)** | 14.34±4.67 | NA | NA |
|  | **PANSS (negative)** | 11.93±4.06 | NA | NA |
|  | **Attention vigilance** |  |  |  |
|  | **Working memory** |  |  |  |
| **FBIRN**  **Site 7** | **Number** | 25 | 27 | NA |
|  | **Age** | 39.56±11.80 | 37.56±10.75 | 0.52 |
|  | **Sex (M/F)** | 20/5 | 20/7 | 0.99 |
|  | **PANSS (positive)** | 16.12±5.41 | NA | NA |
|  | **PANSS (negative)** | 14.04±6.82 | NA | NA |
|  | **Attention vigilance** |  |  |  |
|  | **Working memory** |  |  |  |
| **Total** | **Number** | 151 | 160 | NA |
|  | **Age** | 38.06±11.30 | 37.04±10.68 | 0.41 |
|  | **Sex (M/F)** | 115/36 | 115/45 | 0.99 |
|  | **PANSS (positive)** | 15.32±4.92 | NA | NA |
|  | **PANSS (negative)** | 14.32±5.42 | NA | NA |
|  | **Attention vigilance** |  |  |  |
|  | **Working memory** |  |  |  |

**Supplementary Table 1.**

Demographic and clinical details of subjects for each site of FBIRN dataset.

**Supplementary Table 2.**

The Peak coordinates of components and labels

|  | **Selected Components as Regions of Interest** | **X** | **Y** | **Z** |
| --- | --- | --- | --- | --- |
|  | **Subcortical network (SC)** | | | |
| 1 | Caudate (1) | 6.5 | 10.5 | 5.5 |
| 2 | Subthalamus/hypothalamus (2) | -2.5 | -13.5 | -1.5 |
| 3 | Putamen (3) | -26.5 | 1.5 | -0.5 |
| 4 | Caudate (4) | 21.5 | 10.5 | -3.5 |
| 5 | Thalamus (5) | -12.5 | -18.5 | 11.5 |
|  | **Auditory network (AUD)** | | | |
| 6 | Superior temporal gyrus ([STG], 6) | 62.5 | -22.5 | 7.5 |
| 7 | Middle temporal gyrus ([MTG], 7) | -42.5 | -6.5 | 10.5 |
|  | **Sensorimotor network (SM)** | | | |
| 8 | Postcentral gyrus ([PoCG], 8) | 56.5 | -4.5 | 28.5 |
| 9 | Left postcentral gyrus ([L PoCG], 9) | -38.5 | -22.5 | 56.5 |
| 10 | Paracentral lobule ([ParaCL], 10) | 0.5 | -22.5 | 65.5 |
| 11 | Right postcentral gyrus ([R PoCG], 11) | 38.5 | -19.5 | 55.5 |
| 12 | Superior parietal lobule ([SPL], 12) | -18.5 | -43.5 | 65.5 |
| 13 | Paracentral lobule ([ParaCL], 13) | -18.5 | -9.5 | 56.5 |
| 14 | Precentral gyrus ([PreCG], 14) | -42.5 | -7.5 | 46.5 |
| 15 | Superior parietal lobule ([SPL], 15) | 20.5 | -63.5 | 58.5 |
| 16 | Postcentral gyrus ([PoCG], 16) | -47.5 | -27.5 | 43.5 |
|  | **Visual network (VIS)** | | | |
| 17 | Calcarine gyrus ([CalcarineG], 17) | -12.5 | -66.5 | 8.5 |
| 18 | Middle occipital gyrus ([MOG], 18) | -23.5 | -93.5 | -0.5 |
| 19 | Middle temporal gyrus ([MTG], 19) | 48.5 | -60.5 | 10.5 |
| 20 | Cuneus (20) | 15.5 | -91.5 | 22.5 |
| 21 | Right middle occipital gyrus ([R MOG], 21) | 38.5 | -73.5 | 6.5 |
| 22 | Fusiform gyrus (22) | 29.5 | -42.5 | -12.5 |
| 23 | Inferior occipital gyrus ([IOG], 23) | -36.5 | -76.5 | -4.5 |
| 24 | Lingual gyrus ([LingualG], 24) | -8.5 | -81.5 | -4.5 |
| 25 | Middle temporal gyrus ([MTG], 25) | -44.5 | -57.5 | -7.5 |
|  | **Cognitive-control network (CC)** | | | |
| 26 | Inferior parietal lobule ([IPL], 26) | 45.5 | -61.5 | 43.5 |
| 27 | Insula (27) | -30.5 | 22.5 | -3.5 |
| 28 | Superior medial frontal gyrus ([SMFG], 28) | -0.5 | 50.5 | 29.5 |
| 29 | Inferior frontal gyrus ([IFG], 29) | -48.5 | 34.5 | -0.5 |
| 30 | Right inferior frontal gyrus ([R IFG], 30) | 53.5 | 22.5 | 13.5 |
| 31 | Middle frontal gyrus ([MiFG], 31) | -41.5 | 19.5 | 26.5 |
| 32 | Inferior parietal lobule ([IPL], 32) | -53.5 | -49.5 | 43.5 |
| 33 | Right inferior parietal lobue ([R IPL], 33) | 44.5 | -34.5 | 46.5 |
| 34 | Supplementary motor area ([SMA], 34) | -6.5 | 13.5 | 64.5 |
| 35 | Superior frontal gyrus ([SFG], 35) | -24.5 | 26.5 | 49.5 |
| 36 | Middle frontal gyrus ([MiFG], 36) | 30.5 | 41.5 | 28.5 |
| 37 | Hippocampus ([HiPP], 37) | 23.5 | -9.5 | -16.5 |
| 38 | Left inferior parietal lobule ([L IPL], 38) | -47.5 | 5.5 | 22.5 |
| 39 | Middle cingulate cortex ([MCC], 39) | -15.5 | 20.5 | 37.5 |
| 40 | Inferior frontal gyrus ([IFG], 40) | 39.5 | 44.5 | -0.5 |
| 41 | Middle frontal gyrus ([MiFG], 41) | -26.5 | 47.5 | 5.5 |
| 42 | Hippocampus ([HiPP], 42) | -24.5 | -36.5 | 1.5 |
|  | **Default-mode network (DM)** | | | |
| 43 | Precuneus (43) | -8.5 | -66.5 | 35.5 |
| 44 | Precuneus (44) | -12.5 | -54.5 | 14.5 |
| 45 | Anterior cingulate cortex ([ACC], 45) | -2.5 | 35.5 | 2.5 |
| 46 | Posterior cingulate cortex ([PCC], 46) | -5.5 | -28.5 | 26.5 |
| 47 | Anterior cingulate cortex ([ACC], 47) | -9.5 | 46.5 | -10.5 |
| 48 | Precuneus (48) | -0.5 | -48.5 | 49.5 |
| 49 | Posterior cingulate cortex ([PCC], 49) | -2.5 | 54.5 | 31.5 |
|  | **Cerebellar network (CB)** | | | |
| 50 | Cerebellum ([CB], 50) | -30.5 | -54.5 | -42.5 |
| 51 | Cerebellum ([CB], 51) | -32.5 | -79.5 | -37.5 |
| 52 | Cerebellum ([CB], 52) | 20.5 | -48.5 | -40.5 |
| 53 | Cerebellum ([CB], 53) | 30.5 | -63.5 | -40.5 |

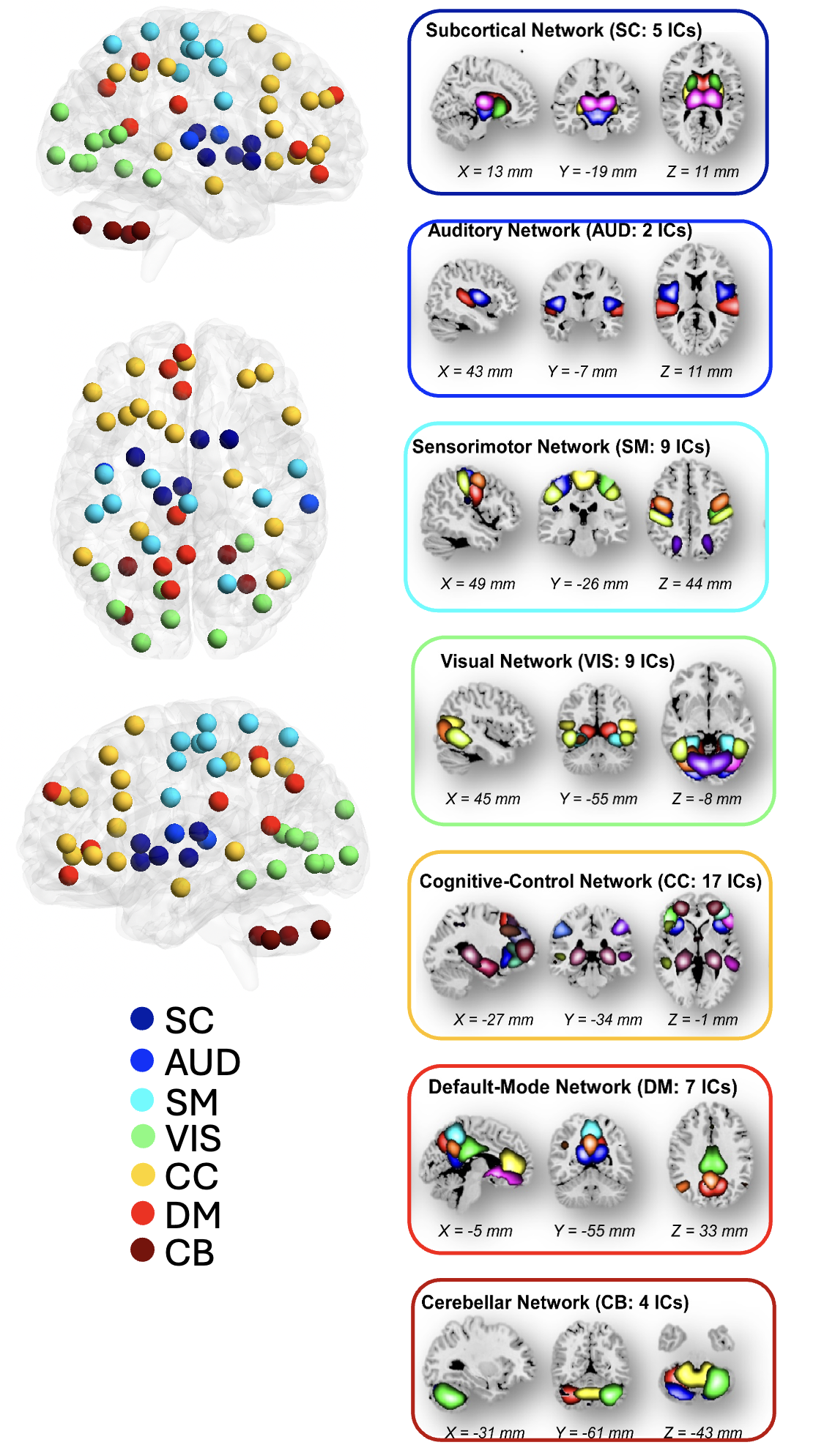

**Supplementary Figure 1.** **The NeuroMark independent component:** We utilized the NeuroMark pipeline to identify reliable intrinsic connectivity networks (ICNs), capturing a total of 53 components that are consistent across independent datasets. These 53 independent components were organized into seven distinct networks: subcortical (SC), auditory (AUD), visual sensory (VSN), sensorimotor (SM), cognitive control (CC), default mode DM), and cerebellar network (CB).

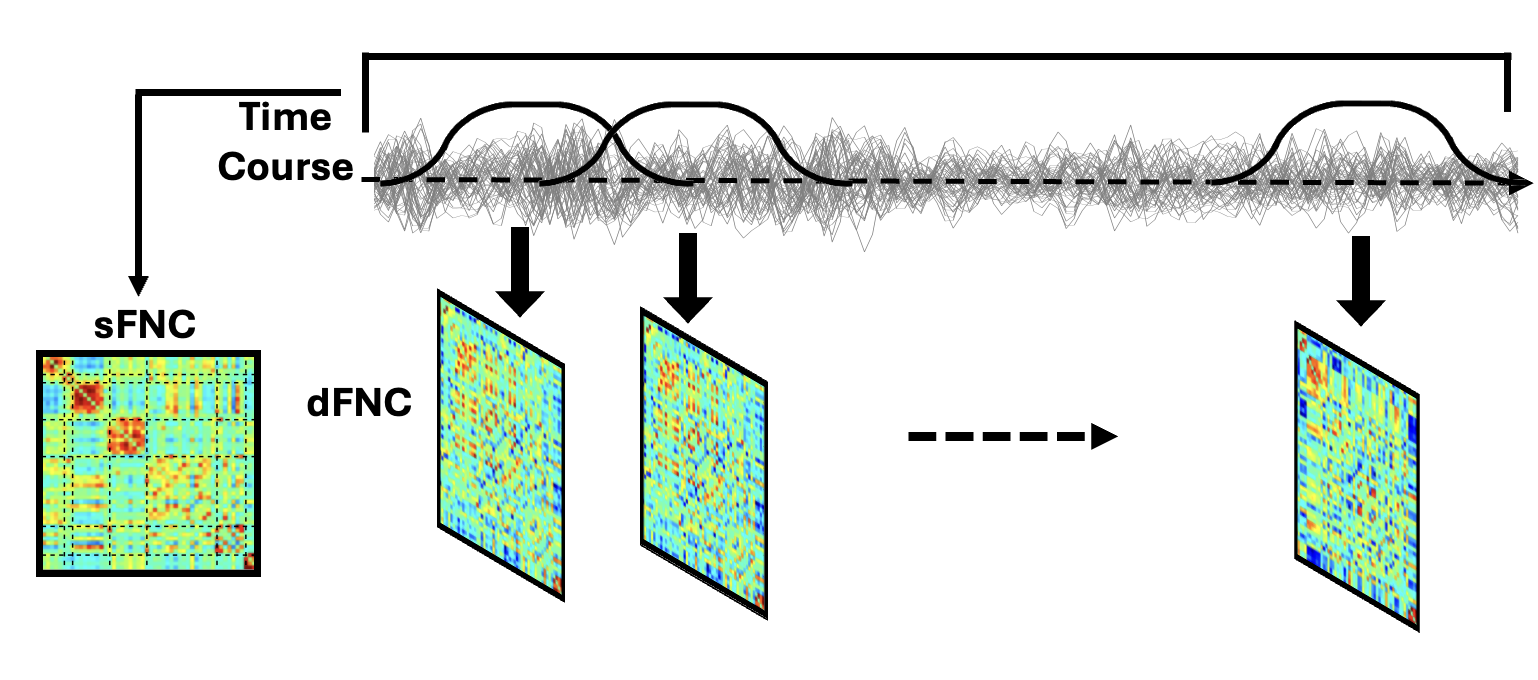

**Supplementary Figure 2.** Static functional network connectivity (FNC) is calculated between any pair of regions across the entire recording period, while dynamic FNC is calculated between pairs of regions for each sliding window throughout the recording.

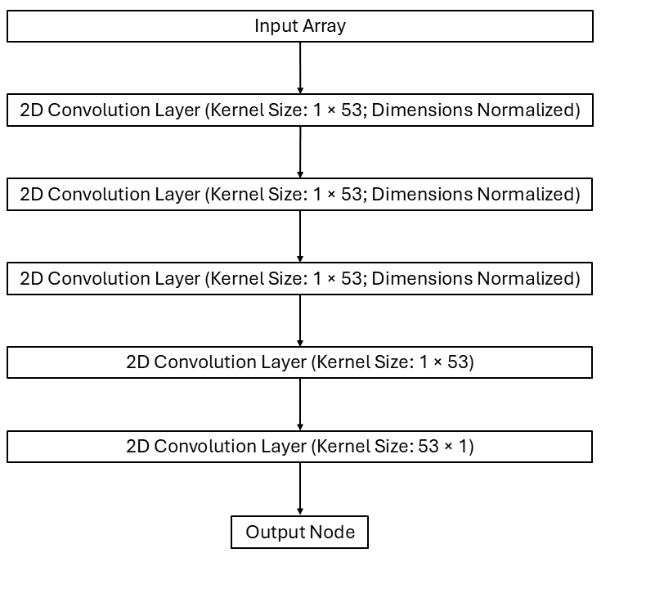

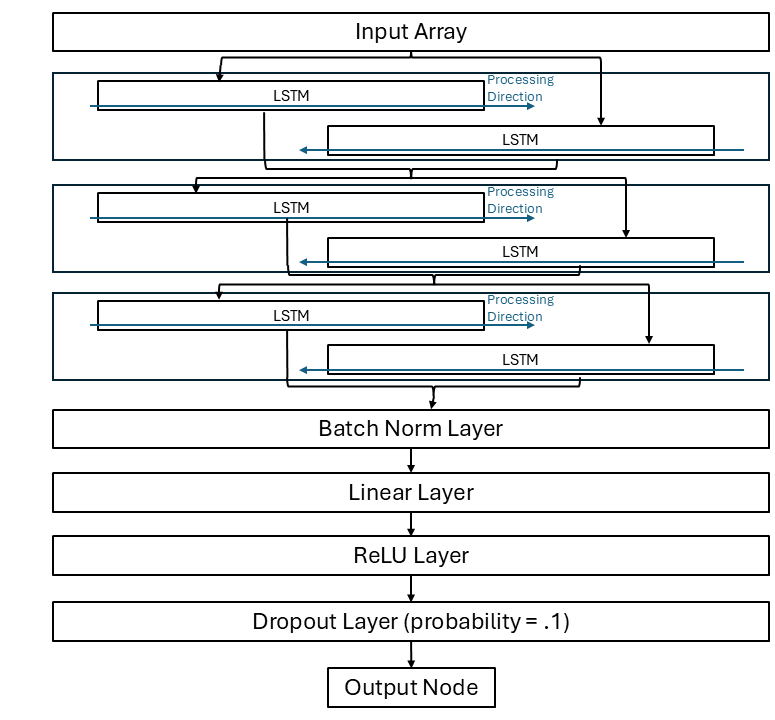

**(B)**

**(B)x**

**Supplementary Figure 3.** **A)** The architecture for the BCGCN model for the sFNC data. It consists of successive convolution layers. The first 3 layers have their dimensions normalized such that the output still consists of 53×53 matrices; these outputs are then used as input to 2 convolution layers which results in each 53×53 matrix becoming a single point. **B)** The architecture for the BiLSTM model for the dFNC data. The first 3 layers are each bidirectional LSTM layers, followed by a Batch Normalization layer, a Linear layer, a ReLU layer, and a Dropout layer (p=0.1), which produces the output.

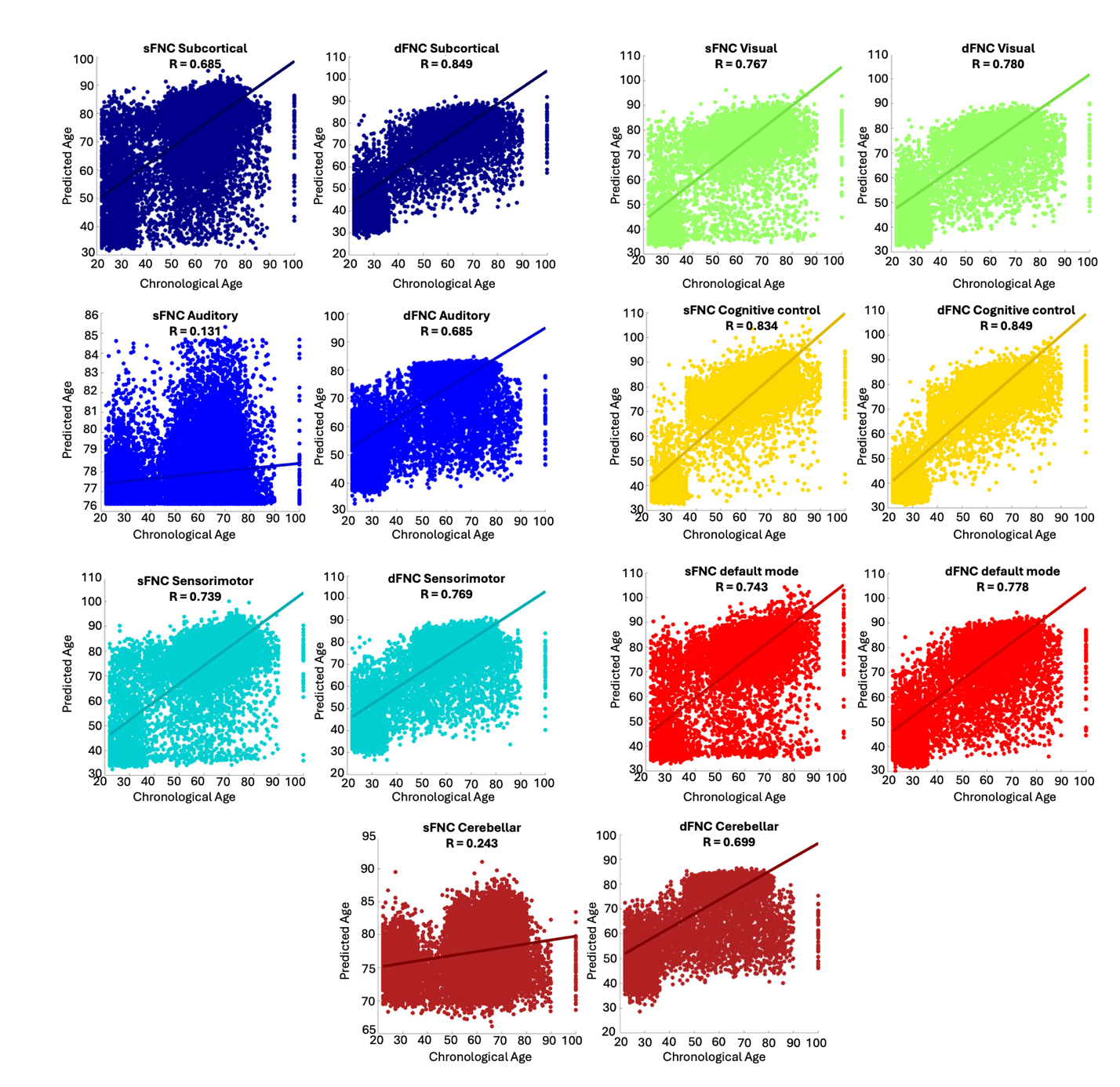

**Supplementary Figure 4.** These graphs depict the individual’s chronological age against their model predicted age for each subnetwork.

**Supplementary Table 3.** Association between subnetwork brain age gap and attention vigilance

| **Network** | **Beta** | **SE** | **R** | **95% CI (lower)** | **95% CI (higher)** | **p value** | **FDR**  **p value** |
| --- | --- | --- | --- | --- | --- | --- | --- |
| **Subcortical (sFNC)** | -0.1813 | 0.0655 | -0.2357 | -0.3111 | -0.0516 | 0.0065 | **0.0228** |
| **Subcortical (dFNC)** | -0.2989 | 0.0896 | -0.2806 | -0.4763 | -0.1215 | 0.0011 | **0.0140** |
| **Auditory (sFNC)** | -0.1086 | 0.0895 | -0.1057 | -0.2858 | 0.0685 | 0.2274 | 0.4548 |
| **Auditory (dFNC)** | -0.0037 | 0.0292 | -0.0112 | -0.0616 | 0.0541 | 0.8977 | 0.9668 |
| **Sensorimotor (sFNC)** | -0.0265 | 0.0285 | -0.0812 | -0.0831 | 0.0299 | 0.3541 | 0.5509 |
| **Sensorimotor (dFNC)** | -0.2153 | 0.0769 | -0.2384 | -0.3675 | -0.0631 | 0.0059 | **0.0228** |
| **Visual (sFNC)** | -0.0217 | 0.0224 | -0.0846 | -0.0661 | 0.0226 | 0.3347 | 0.5509 |
| **Visual (dFNC)** | -0.0061 | 0.0245 | -0.0218 | -0.0546 | 0.0423 | 0.8032 | 0.9668 |
| **Cognitive control (sFNC)** | -0.0353 | 0.0145 | -0.2081 | -0.0641 | -0.0065 | 0.0166 | **0.0465** |
| **Cognitive control (dFNC)** | -0.2928 | 0.1523 | -0.1662 | -0.5942 | 0.0086 | 0.0568 | 0.1326 |
| **Default mode (sFNC)** | 0.0007 | 0.0237 | 0.0026 | -0.0463 | 0.0477 | 0.9758 | 0.9758 |
| **Default mode (dFNC)** | -0.1053 | 0.0333 | -0.2666 | -0.1713 | -0.0392 | 0.0020 | **0.0140** |
| **Cerebellar (sFNC)** | 0.0285 | 0.0520 | 0.0479 | -0.0744 | 0.1315 | 0.5849 | 0.8188 |
| **Cerebellar (dFNC)** | -0.0181 | 0.0838 | -0.0189 | -0.1840 | 0.1477 | 0.8288 | 0.9668 |

**Supplementary Table 4.** Association between subnetwork brain age gap and working memory

| **Network** | **Beta** | **SE** | **R** | **95% CI (lower)** | **95% CI (higher)** | **p value** | **FDR**  **p value** |
| --- | --- | --- | --- | --- | --- | --- | --- |
| **Subcortical (sFNC)** | -0.1050 | 0.0518 | -0.1724 | -0.2076 | -0.0020 | 0.0447 | 0.2088 |
| **Subcortical (dFNC)** | -0.1473 | 0.0714 | -0.1754 | -0.2885 | -0.0060 | 0.0410 | 0.2088 |
| **Auditory (sFNC)** | -0.1069 | 0.0699 | -0.1309 | -0.2453 | 0.0313 | 0.1286 | 0.3272 |
| **Auditory (dFNC)** | -0.0123 | 0.0227 | -0.0467 | -0.0572 | 0.0326 | 0.5888 | 0.6341 |
| **Sensorimotor (sFNC)** | -0.0187 | 0.0223 | -0.0721 | -0.0630 | 0.0255 | 0.4037 | 0.5139 |
| **Sensorimotor (dFNC)** | -0.0799 | 0.0602 | -0.1138 | -0.1992 | 0.0392 | 0.1870 | 0.3272 |
| **Visual (sFNC)** | -0.0199 | 0.0172 | -0.0990 | -0.0540 | 0.0142 | 0.2510 | 0.3514 |
| **Visual (dFNC)** | -0.0249 | 0.0187 | -0.1138 | -0.0620 | 0.0122 | 0.1869 | 0.3272 |
| **Cognitive control (sFNC)** | -0.0136 | 0.0115 | -0.1010 | -0.0365 | 0.0092 | 0.2417 | 0.3514 |
| **Cognitive control (dFNC)** | -0.1639 | 0.1196 | -0.1174 | -0.4006 | 0.0728 | 0.1731 | 0.3272 |
| **Default mode (sFNC)** | 0.0331 | 0.0183 | 0.1540 | -0.0031 | 0.0694 | 0.0734 | 0.2569 |
| **Default mode (dFNC)** | -0.0662 | 0.0264 | -0.2114 | -0.1186 | -0.0139 | 0.0134 | 0.1884 |
| **Cerebellar (sFNC)** | 0.0226 | 0.0405 | 0.0482 | -0.0574 | 0.1027 | 0.5766 | 0.6341 |
| **Cerebellar (dFNC)** | 0.0286 | 0.0648 | 0.0380 | -0.0997 | 0.1569 | 0.6598 | 0.6598 |
